# Association of *HLA-DRB1* alleles with rheumatoid arthritis in Mediterranean populations: A meta-analysis with regional differences, advanced heterogeneity and evidence quality assessment

**DOI:** 10.1101/2025.04.29.25326652

**Authors:** Jamil Mourad, Sarah Naji, AbdulKarim El Karaaoui, Doha Wahbeh

**Author notes:** Corresponding author: (JM).

## Abstract

**Objective:** This meta-analysis evaluates the association between *HLA-DRB1* alleles and rheumatoid arthritis (RA) susceptibility in Mediterranean populations, incorporating regional subgroup analyses, heterogeneity assessment, and evidence quality grading.

**Methods:** A systematic search was conducted across 11 major databases and grey literature sources until December 31, 2023. Eligible studies reported *HLA-DRB1* allele frequencies in RA patients and controls from Mediterranean countries. Pooled odds ratios were calculated using random-effects models. Heterogeneity was assessed using Cochrane’s Q, I², prediction intervals, cumulative distribution function and meta-regression. Risk of bias was evaluated using the Risk of Bias in Non-randomized Studies—of Exposures (ROBINS-E) tool. Publication bias was examined using funnel-plots, the trim-and-fill method, and Rücker’s-test. Sensitivity analyses excluded studies with very-high risk of bias and performed leave-one-out diagnostics. Post hoc statistical power analyses were performed for each allele comparison. Evidence certainty was evaluated using the Grading of Recommendations Assessment, Development, and Evaluation (GRADE) framework. Registered in PROSPERO (CRD42025641117).

**Results:** Forty-eight studies from 13 Mediterranean countries (6,536 RA cases, 10,211 controls) were included. *HLA-DRB1*\*01, *04, *09, and *10 were associated with increased RA risk, with *04 showing the highest pooled effect size (OR = 2.412, 95%CI [2.204–2.641]). *HLA-DRB1*\*03, *07, *08, *11, *12, *13, and *14 were protective, with *13 having the lowest odds ratio (OR = 0.580, 95%CI [0.527–0.638]). Subgroup analyses identified additional risk variants (*01:01, *04:01, *04:04, *04:05, *04:08, and *10:01) and one protective allele (*04:03). No significant regional differences were found between Southern Europe and the Middle East and North Africa. Heterogeneity ranged from moderate to high (I² = 37.9%– 61.1%). Most allele comparisons exhibited sufficient power, although some associations (*03, *08, *09, and *04:03) were unstable in sensitivity analyses. Overall certainty of evidence was rated low to very low.

**Conclusions:** HLA-DRB1*04 and *10 were strongly associated with RA risk, while *13 and *07 were protective. Although HLA-DRB1*09 was significantly associated with RA, it was unstable and supported by very low quality evidence. Despite similar patterns across regions, heterogeneity and study limitations underscore the need for better-controlled, population-specific genetic studies.

## Introduction

Rheumatoid arthritis (RA) is a chronic autoimmune disease affecting 0.5–1% of the global population, with women being 2–3 times more susceptible than men [1–3]. RA prevalence varies geographically, with higher rates in industrialized regions (0.35–0.38%) and lower rates in Oceania (0.14%) and sub-Saharan Africa (0.13%) [4]. In Mediterranean countries (Southern Europe, Middle East, and North Africa), intermediate rates of 0.12–0.3% have been reported, influenced by unique genetic and environmental factors [5].

RA prevalence reflects the chronic disease burden, whereas regional variations in incidence rates highlight differences in environmental exposures and diagnostic practices [6]. The MENA (Middle East and North Africa) report the highest incidence (37 per 100,000 patient-years), followed by North America (22.5 per 100,000) and Western Europe (20–30 per 100,000). In contrast, lower rates are observed in Southeast Asia (6.2), Oceania (16), and Latin America (18.5) per 100,000 patient-years [4].

*HLA-DRB1* alleles, located on chromosome 6p21.3 within the major histocompatibility complex (MHC), are critical genetic contributors [7, 8]. These variants are pivotal in antigen presentation and T-cell activation, which is central to RA pathogenesis. Specific *HLA-DRB1* alleles containing the shared epitope (SE) motif (*HLA-DRB1*\*01:01, *04:01, *04:04, *04:05, *10:01, and *14:02) are strongly associated with increased susceptibility to RA [8]. The SE motif, a conserved sequence at positions 70-74, binds citrullinated peptides vital to disease mechanisms [9]. Furthermore, the *HLA-DRB1*\*09 allele, although lacking the SE motif, has been linked to a heightened risk of RA in East Asian populations [10], highlighting the complex genetic landscape of this disease.

Beyond the SE motif, studies have identified amino acid variations at positions 11, 71, and 74 of the *HLA-DRB1* molecule as key determinants of RA risk. These residues are integral to peptide binding and strongly influence disease susceptibility [11].

Conversely, some *HLA-DRB1* alleles are protective against RA. These alleles include *HLA-DRB1*\*01:03, *04:02, *11:02, *11:03, *13:01, *13:02, and *13:04, which possess the *DERAA* sequence at positions 70-74, conferring resistance to RA development [12]. Additionally, classifications such as S1, S2, S3P, and S3D groups have been proposed to refine risk stratification. These classifications account for amino acid changes that modulate the protective or predisposing effects of the SE motif [13, 14].

Geographic and genetic variability in *HLA-DRB1* allele distribution contributes to differences in RA susceptibility globally [5, 15]. While environmental factors, such as the Mediterranean diet rich in anti-inflammatory components (e.g., olive oil, fish), smoking rates, and varying infectious exposures, contribute to RA risk and disease course in Mediterranean populations [6, 16], these factors alone cannot fully explain RA patterns. Understanding the role of hereditary contributors remains critical, particularly in this historically interconnected and environmentally diverse region.

Despite extensive research on *HLA-DRB1* alleles, their role in RA susceptibility across the Mediterranean region remains insufficiently characterized. Previous meta-analyses have primarily focused on Southern Europe [17], which restricts the applicability of their findings to the broader regional populations. This fragmented approach overlooks the shared genetic, historical, and geo-environmental interactions that have influenced allele distributions within the Mediterranean basin over centuries [18]. Furthermore, previous meta-analyses have not incorporated heterogeneity analyses to identify the underlying sources of variability. Additionally, the lack of systematic evidence quality assessment has weakened the reliability and generalizability of their findings.

This study aims to conduct the first comprehensive meta-analysis assessing *HLA-DRB1* alleles in RA susceptibility across the Mediterranean region, with a comparative analysis between Southern Europe and the MENA region. This research effort systematically examines risk and protective *HLA-DRB1* alleles, incorporates advanced heterogeneity analyses, and evaluates the quality of existing evidence, providing new insights into the role of *HLA-DRB1* in RA susceptibility within Mediterranean populations. These findings have important implications for region-specific genetic screening and the development of tailored therapeutic strategies.

## Materials and methods

### General methodology

All methodological steps, including study selection, data extraction, risk of bias assessment, and quality grading, were conducted independently by two reviewers, with discrepancies resolved by discussion or consultation with a third reviewer. The systematic review and meta-analysis adhered to PRISMA 2020 guidelines [19] (**S1 *Checklist***), the PRISMA Abstract Checklist (**S2 Checklist**), and the Genetic Association Checklist (**S3 Checklist**) in accordance with PLOS ONE requirements. The study was prospectively registered in PROSPERO (CRD42025641117), and the full protocol, including amendments, is publicly available at: https://www.crd.york.ac.uk/PROSPERO/view/CRD42025641117.

**Ethics statement**: No approval was required as this meta-analysis used published data.

#### Study selection

The study selection process followed a structured and transparent approach in accordance with the PRISMA guidelines [19].

#### Systematic search

A thorough search of the literature was performed across several databases, including PubMed, Embase, Web of Science, Scopus, Index Medicus, Google Scholar, Cochrane Library, Scielo, Theses.fr, and WorldCat. These resources were chosen to ensure a wide-ranging overview of biomedical studies while also incorporating multidisciplinary indexing and journals specific to different regions, as well as enabling comprehensive searches that include grey literature. The search encompassed all available records up until December 31, 2023, and were conducted between August and December 2024.

Researchers used the keywords “Rheumatoid Arthritis”, “*HLA*” and “*HLA-DRB1*” using Boolean operators to find relevant studies. Tailored search strategies specific to each database were implemented including:

- **PubMed**: ((Rheumatoid Arthritis [Title/Abstract]) AND (HLA[Title/Abstract])) AND (DRB1[Title/Abstract])
- **Embase**: (Rheumatoid Arthritis and HLA).ti
- **Web of Science**: (TI=(Rheumatoid Arthritis)) AND (TI=(HLA)) Timespan: 1990-01-01 to 2023-12-31
- **Scopus**: (TITLE(rheumatoid AND arthritis) AND TITLE(HLA)) AND PUBYEAR > 1990 AND PUBYEAR <= 2023
- **Index Medicus**: (ti:(Rheumatoid Arthritis)) AND (ti:(HLA))
- **Google Scholar**: allintitle: “HLA rheumatoid arthritis”
- **Cochrane Library**: (”rheumatoid arthritis” in Title Abstract Keyword) AND (”HLA” in Title Abstract Keyword)
- **Scielo**: (ti:(Rheumatoid Arthritis)) AND (ti:(HLA))
- **Theses.fr**: titres.*:(Rheumatoid Arthritis) AND titres.*:(HLA)
- **WorldCat**: ti:HLA AND ti:Rheumatoid Arthritis (limited to theses and dissertations, publication years 1990–2023).

To minimize the risk of publication bias and to ensure we captured a range of unpublished or lesser-known studies, we included grey literature sources such as WorldCat and Theses.fr. Additionally, we performed a thorough screening of Google Scholar to account for broader and less specific indexing.

#### Screening and eligibility

Search results from all databases were first exported to EndNote X9 (Thomson Reuters, 2018) for initial organization. The cleaned dataset was then exported as an XML file and uploaded to Covidence.org [20] for systematic screening where duplicate entries have been automatically removed. Remaining duplicates were reviewed and removed in the title and abstract screening phase.

Studies were included if they had an observational design (case-control or cohort) with cases and controls matched by gender and nationality to reduce confounding. Eligible studies reported either allele frequencies (AF) or allele counts (AC) for *HLA-DRB1* in RA patients and controls. Rheumatoid arthritis had to be diagnosed based on at least one of the following: the 1987 ACR classification criteria, the 2010 ACR/EULAR criteria, or clinical features indicative of RA when formal classification criteria were not provided. Only studies conducted in Mediterranean countries were considered.

Exclusion criteria included studies conducted outside the Mediterranean region, studies lacking a control group or retrievable *HLA-DRB1* data, and studies with overlapping populations (only the most comprehensive dataset was retained). Conference abstracts, case reports, and editorials were excluded unless full data were available.

To enhance coverage, authors of conference abstracts and unretrievable articles were contacted via email for full texts when needed. Reference lists of all relevant studies were manually screened for additional articles. Non-English studies were translated, and translations were verified for accuracy and reliability.

### Data extraction

First author’s name, year of publication, country of study, number of cases and controls, *HLA-DRB1* alleles for cases and controls, diagnostic method and *HLA* genotyping method where systematically extracted from each study.

For studies reporting allele frequencies, allele counts for cases and controls were calculated using standard conversion formulas to harmonize data across studies. Extraction followed independent review with conflict resolution, as detailed in the General methodology section. All extracted data were cross verified to minimize transcription errors and ensure accuracy.

### Risk of bias

The risk of bias in the included studies was assessed independently using Cochrane’s Risk of Bias in Non-randomized Studies of Exposures (ROBINS-E) tool [21]. The ROBINS-E tool evaluates seven domains of bias, including confounding factors, participant selection, exposure classification, deviations from intended interventions, missing data, outcome measurement, and reported results selection. For this meta-analysis, particular attention was given to confounding factors (age, gender, ethnicity, smoking, and diet) [22] and participant selection, as these were deemed most critical to study validity.

Each study was categorized into one of four levels of overall risk: low, some concerns, high, or very high.

A traffic light plot was generated in R [23] to visually represent domain-specific risk of bias across studies, aiding in the assessment of overall study credibility.

### Statistical analysis

Meta-analyses were conducted for all *HLA-DRB1* variants supported by data from at least nine independent studies, allowing for reliable pooled estimates and meaningful heterogeneity assessment. Odds ratios (ORs) and 95% confidence intervals (CIs) were calculated using a random-effects model [24]. Statistical significance was evaluated using the z-test on the pooled log (OR), with *p*-values below 0.05 considered significant.

All analyses were performed by applying the DerSimonian-Laird random-effects model to account for between-study variability. This approach was chosen given the expected differences in study populations and methodological characteristics [25].

The overall risk of bias (ROB) for each meta-analysis was calculated using a weighted average approach, where each study’s ROB score was adjusted based on its relative contribution to the pooled estimate. To achieve this, ROB ratings from ROBINS-E were converted into numerical values (Low = 1, Some Concerns = 2, High = 3, Very High = 4), and study weights (wᵢ) were assigned using the inverse variance method (1/SE²) [24, 25]. The overall ROB was computed as:

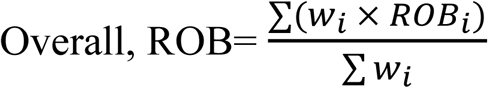

Interpretation thresholds were: ≤1.5 = Low Risk, >1.5 – 2.5 = Some Concerns, >2.5 – 3.5 = High Risk, >3.5 = Very High Risk. This approach improves transparency by ensuring that studies with greater statistical power contribute more to the ROB assessment, enhancing the reliability of bias evaluation. It should be noted that this scoring and weighting framework was developed specifically for this review and is not part of the original ROBINS-E or GRADE (Grading of Recommendations, Assessment, Development and Evaluation) guidance [21, 26]

#### Data management and software

All data were compiled and preprocessed using Microsoft^®^ Office Excel^®^ to enable standardization and facilitate manual validation. Statistical analyses, including pooled effect size calculations, overall risk of bias estimation, heterogeneity assessments, forest plots, and funnel plots, were conducted using the ***meta*** package in R. Advanced analyses, such as heterogeneity analyses, sensitivity analyses and outlier detection, were performed using the ***dmetar*** package in R [27].

#### Post hoc power calculation

For each *HLA-DRB1* allele meta-analysis, post hoc statistical power was computed using a two-sample z-test for proportions with unequal group sizes. Effect sizes were calculated as Cohen’s h, defined as the difference between the arcsine-transformed square roots of allele frequencies in RA cases and controls. Power estimates were derived from observed allele frequencies and total sample sizes, assuming a two-sided alpha of 0.05 [28]. Analyses were conducted in R using custom scripts and base functions from the ***stats*** and ***dplyr*** packages [23]. A power estimate ≥80% was considered high; values below this threshold were classified as low.

### Heterogeneity analysis

Heterogeneity across the meta-analyses was assessed using multiple statistical methods to capture variability among the included studies. Cochran’s Q-statistics were employed to determine whether observed variability in effect sizes exceeded what would be expected by chance, though its interpretation can be influenced by the number of studies and their sample sizes. The I² statistic quantified the proportion of variability attributable to true heterogeneity rather than sampling error. Prediction Intervals (PI) were calculated to estimate the range within which the true effect sizes of 95% of comparable populations are likely to fall, offering additional insights into the variability not captured by Q-statistic or I² alone [29].

To assess whether geographic region accounted for observed heterogeneity, meta-regression was conducted using the ***rma*** function from the ***metafor*** package in R. The model included region (Southern Europe vs. MENA) as a moderator and was fitted using restricted maximum likelihood (REML). Odds ratios were log-transformed, and inverse-variance weights were applied. Statistical significance was set at *p* < 0.05. Diagnostic plots were generated to evaluate model fit and residual heterogeneity [30].

Outliers, defined as studies with effect sizes significantly deviating from the pooled estimate, were identified and removed using the ***find.outliers*** function in the **dmetar** package in R [27]. Their removal assessed the extent to which individual studies influenced heterogeneity and overall estimates.

For meta-analyses with persistent heterogeneity after outlier removal, we estimated the percentage of true effect sizes expected to fall below or above an odds ratio (OR) of 1 using Prediction Intervals. This involved log-transforming the OR and the bounds of the PI, estimating the standard deviation, and applying the cumulative distribution function (CDF) of the standard normal distribution via the ***pnorm*** function in R [31, 32]. These calculations, based on methodologies outlined by Borenstein (2024), helped contextualize the variability in effect sizes across studies [33].

### Publication bias

Two methods were used to assess publication bias in this meta-analysis, either the Trim-and-Fill method or the Rucker’s test, depending on the level of heterogeneity of the included datasets.

When substantial heterogeneity was observed, the possibility of publication bias was evaluated using the trim-and-fill method, which estimates the number of potentially missing studies and adjusts the pooled effect size accordingly. This analysis was conducted using the ***trimfill*** function from the ***dmetar*** package in R [27]. Funnel plot asymmetry was assessed visually, and the difference between the original and adjusted pooled effect sizes was examined to gauge the potential impact of publication bias [34]. Unlike Egger’s test, which assumes that funnel plot asymmetry arises solely due to publication bias, the trim-and-fill method is less likely to yield misleading results in the presence of high heterogeneity [34, 35].

Publication bias was classified based on the number of imputed studies using the trim-and-fill method: minimal bias (up to 3 missing studies), moderate bias (4–5 studies), and substantial bias (6 or more studies). These thresholds were defined as a priori for the purpose of this analysis.

For datasets exhibiting no detectable heterogeneity and symmetrical distribution of effect sizes in funnel plots, Rücker’s test was applied. This method is considered more appropriate than Egger’s test under low heterogeneity conditions, as it offers greater sensitivity to funnel plot asymmetry while maintaining a lower false positive rate [36, 37]. A *p*-value less than 0.1 was considered indicative of potential publication bias.

### Sensitivity analysis

To evaluate the stability of the meta-analysis findings, several sensitivity analyses were conducted. First, studies classified as having a very high risk of bias (per the ROBINS-E tool) were excluded to assess their influence on the pooled estimates. Changes in effect size following their removal were examined to determine whether the overall results were dependent on lower-quality studies.

Next, a leave-one-out procedure was applied, systematically omitting one study at a time to evaluate the impact of individual studies on the summary effect. This approach helped identify studies with disproportionate influence, ensuring that no single dataset unduly shaped the overall results [38].

Finally, the trim-and-fill method was used to explore the potential effect of publication bias. This technique estimated the number of potentially missing studies and recalculated the pooled effect after imputing them. A substantial shift between the original and adjusted estimates was interpreted as evidence of publication-related distortion [39].

### Quality of evidence assessment

The certainty of evidence for each outcome was evaluated using the GRADE framework. As all included studies were observational, each outcome began with a baseline rating of low certainty. Certainty was subsequently downgraded or upgraded based on five core domains: risk of bias, inconsistency, indirectness, imprecision, and publication bias. Upgrades were applied in cases of large effect sizes or when plausible confounding was well-controlled [26, 40].

#### Downgrade Criteria

- Risk of Bias: Downgraded by one level for moderate concerns (some concerns in ROBINS-E) and by two levels for serious concerns (high risk).
- Inconsistency: Downgraded by one level if heterogeneity was moderate (I² = 30– 60%) and by two levels if substantial (I² > 60%).
- Imprecision: Downgraded if confidence intervals were narrow but centered near the null (within ±0.1 of OR = 1), or if they were wide and encompassed both potential harm and benefit.
- Indirectness: Downgraded when differences in the population, intervention, or outcome limited applicability to the research question.
- Publication Bias: Downgraded based on evidence of funnel plot asymmetry and the number of studies imputed by the trim-and-fill method (one level for moderate bias: 3–5 studies; two levels for serious bias: ≥6 studies).

#### Upgrade Criteria

- Large Effect: Upgraded by one level for a strong association (OR > 2.0 or < 0.5) and by two levels for a very strong association (OR > 5.0 or < 0.2).
- Control of Confounders: Upgraded by one level if plausible confounding factors were adequately addressed and the effect estimate remained reliable.

#### Sequential Adjustment

Downgrades were applied first, with one level for serious concerns and two levels for very serious concerns across domains. If an outcome received two or more downgrades, it was classified as very low certainty, and no further downgrades were applied. Upgrades were applied subsequently, with each strong positive factor increasing certainty by one level (to a maximum of two levels). If multiple downgrades resulted in an overall certainty rating of very low, this rating was retained even if upgrading criteria were later met.

#### Final Categorization

The final certainty of evidence was classified as very low, low, moderate, or high, based on the net balance of downgrades and upgrades applied to each outcome [26, 40].

## Results

### Literature search

A comprehensive systematic search identified a total of 6,412 records across multiple databases. After removing duplicates, screening titles and abstracts, and reviewing full texts, 102 articles initially met the inclusion criteria. Following detailed eligibility assessment, 48 studies were ultimately included in the meta-analysis. Studies were excluded after full-text review due to reasons such as wrong exposure, insufficient data for meta-analysis, or inability to retrieve the full text. A detailed list of excluded full-text studies, including authors, titles, and reasons for exclusion, is provided in (S1 ***Checklist-PRISMA 2020 Checklist***

**S2 Checklist-PRISMA Abstract Checklist**

**S3 Checklist-Genetic Association Meta-analysis Checklist**

**S4** Table).The complete study selection process is illustrated in the PRISMA 2020 flow diagram (**Fig 1**).

**Fig 1:**
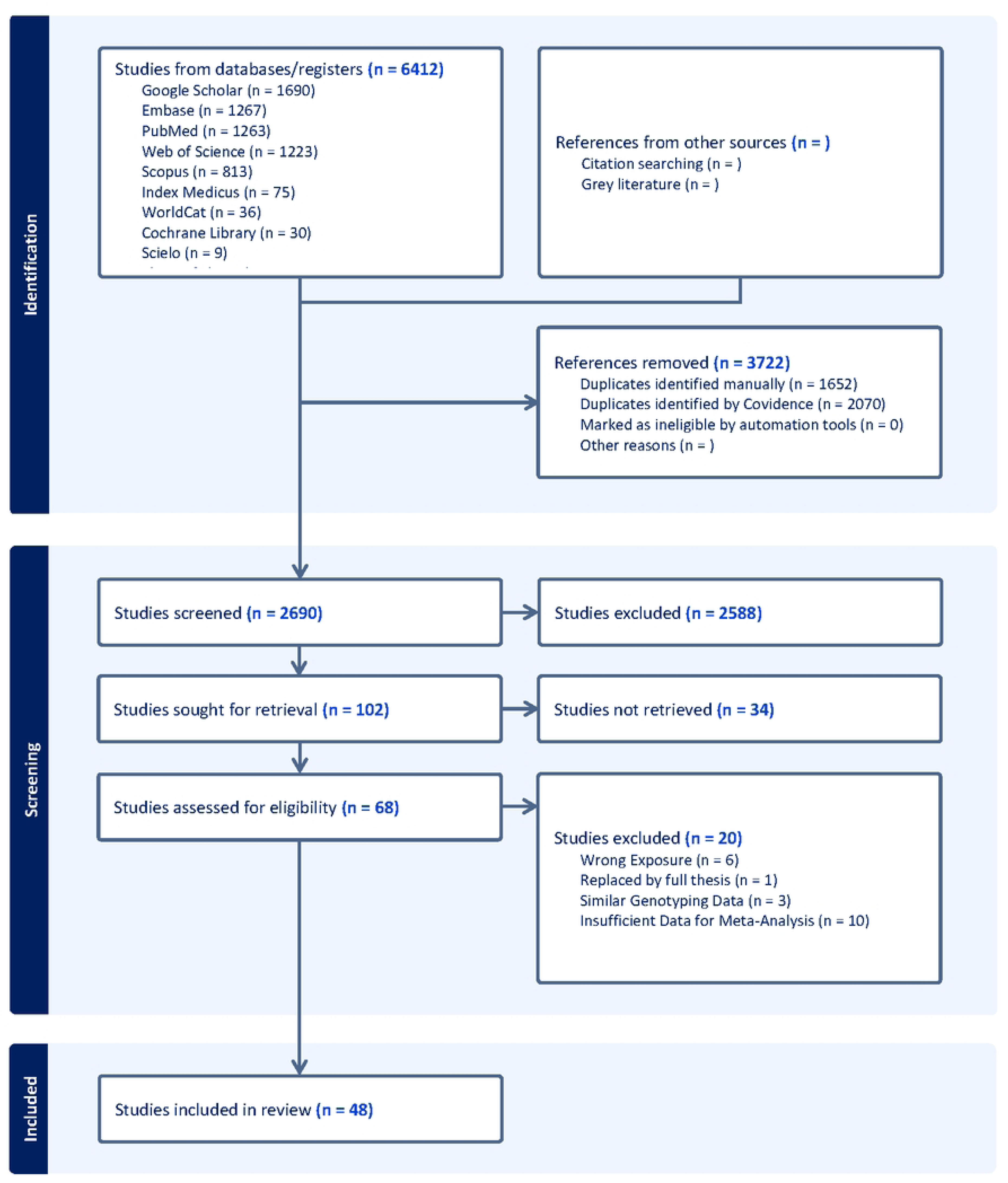
PRISMA 2020 flow diagram representing the study selection process for inclusion in the meta-analysis. The figure was generated using Covidence.org and was manually revised to accurately reflect the status of full-text retrieval and to specify grey literature sources.

**Fig 2.**
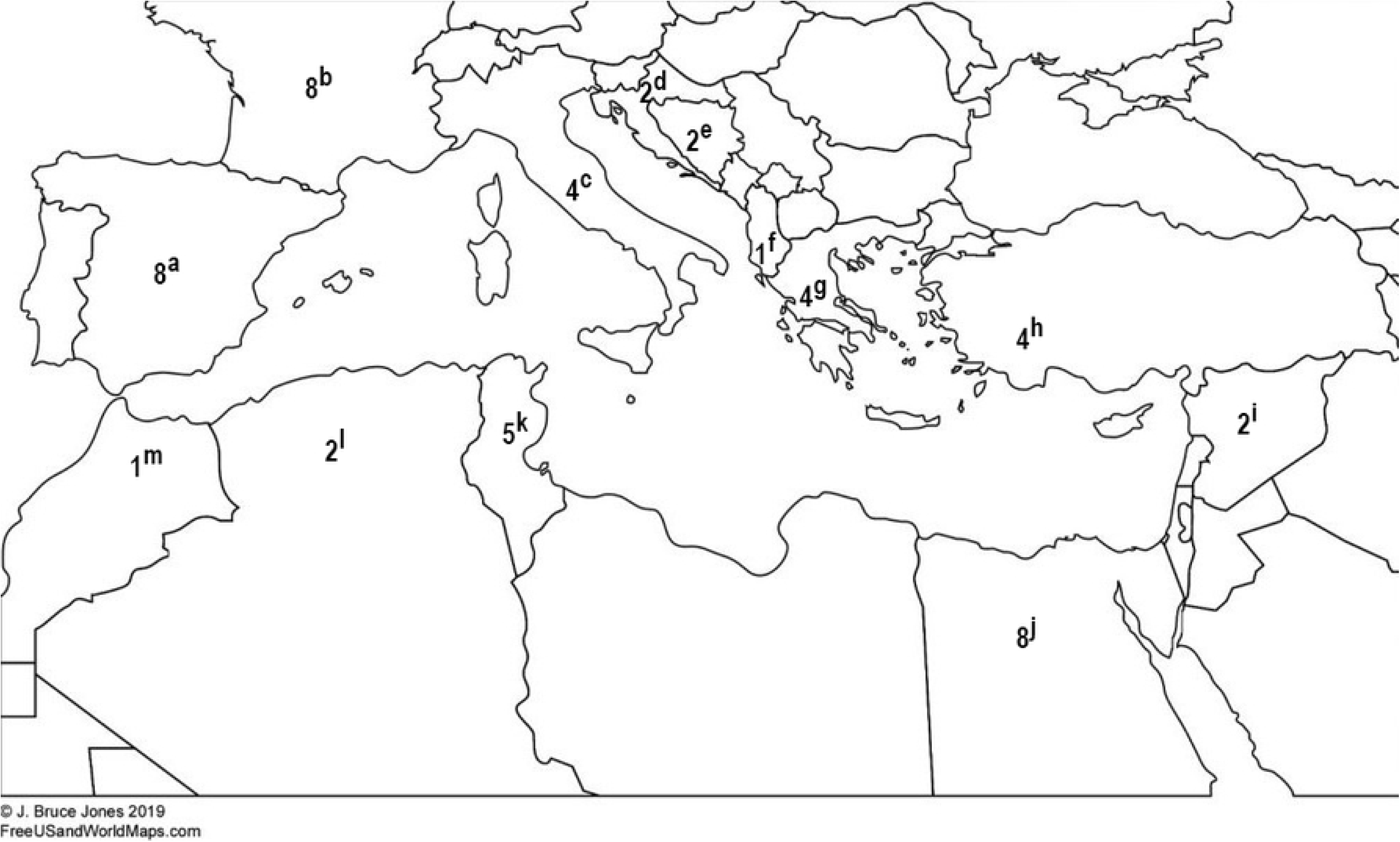
Number and location of studied populations. Letters represent: (a) Spain; (b) France; (c) Italy; (d) Croatia; (e) Bosnia and Herzegovina; (f) Albania; (g) Greece; (h) Turkey; (i) Syria; (j) Egypt; (k) Tunisia; (l) Algeria; (m) Morocco. The base map was adapted from a royalty-free image by J. Bruce Jones, 2019 (FreeUSandWorldMaps.com) and modified to display the number of studies contributed by each country.

To retrieve inaccessible full texts and conference abstracts, authors were contacted via email. Three authors responded:

- The author of a conference abstract [41] shared her thesis [42].
- The author of a conference abstract [43] provided the full text of her published article [44], which was already included in our database.
- The author of a similar meta-analysis [45] supplied the full text of an article [46], which was included in our database and met the inclusion criteria.

These contributions ensured the inclusion of critical data, enhancing the comprehensiveness of the analysis.

### Study characteristics

The meta-analysis included data from 48 studies representing 13 Mediterranean countries, with a total of 6,536 RA patients (26–857 per study) and 10,211 controls (14–2,178 per study). Among patients, the proportion of women ranged from 39% to 100%, while controls ranged from 25% to 100%. The mean age of RA patients spanned 35.4 to 64 years, compared to 27.3 to 61.3 years in controls. Disease duration among RA patients averaged 2 to 25 years.

Three HLA typing techniques were employed across the studies: Polymerase Chain Reaction - Single Specific Primer (PCR-SSP); Polymerase Chain Reaction - Sequence-Specific Oligonucleotide (PCR-SSO); Reverse Dot-Blot (RDB) Hybridization.

Most studies utilized case-control design and relied on the 1987 ACR or 2010 EULAR/ACR diagnostic criteria for RA. Two studies contributed data from distinct subpopulations [47, 48]. **Table 1** provides a comprehensive summary of study characteristics, while ( **2**) illustrates the geographic distribution of included studies.

**Table 1:** Characteristics of Studies Included in the Meta-Analysis of *HLA-DRB1* Alleles and Rheumatoid Arthritis in Mediterranean Populations:

|  | **Study**<br>**1st Author; Year** | **Reference** | **Study**<br>**Design** | **Country** | **HLA Typing** | **Data**<br>**Form** | **Cases**<br>**(n)** | **Controls**<br>**(n)** | **Diagnosis**<br>**Criteria** |
| --- | --- | --- | --- | --- | --- | --- | --- | --- | --- |
| **1** | Abel El-wahed et al; 2018 | **[49]** | Case Control | Egypt | PCR-SSO | AC | 48 | 48 | 1987 ACR |
| **2** | Acheli et al; 2014 | **[42]** | Case Control | Algeria | PCR-SSP | AC | 159 | 110 | 1987 ACR |
| **3** | Angellini et al; 1992 | **[50]** | Case Control | Italy | PCR-SSO | AC | 48 | 109 | 1987 ACR |
| **4** | Atouf et al; 2008 | **[51]** | Case Control | Morroco | PCR-SSP | AC | 49 | 183 | 1987 ACR |
| **5** | Balandraud et al; 2013 | **[52]** | Cohort | France | PCR-SSO | AC | 857 | 2178 | 2010 EULAR/ACR |
| **6** | Balsa et al; 2010 | **[53]** | Cohort | Spain | PCR-SSO | AC | 253 | 173 | 1987 ACR |
| **7** | Batmaz et al; 2013 | **[54]** | Case Control | Turkey | PCR-SSP | AC | 96 | 84 | 1987 ACR |
| **8** | Ben Hamad et al; 2012 | **[55]** | Case Control | Tunisia | PCR-SSP | AF | 142 | 123 | 1987 ACR |
| **9** | Benazet et al; 1995 | **[56]** | Case Control | France | PCR-SSO | AC | 73 | 108 | 1987 ACR |
| **10** | Beri et al; 2005 | **[57]** | Case Control | Italy | PCR-SSO | AC | 101 | 229 | 1987 ACR |
| **11** | Boki et al; 1992 | **[58]** | Case Control | Greece | PCR-SSO | AC | 92 | 84 | 1987 ACR |
| **12** | Bonji et al; 2004 | **[59]** | Case Control | Italy | PCR-SSO | AC | 264 | 532 | 1987 ACR |
| **13** | Boussaid et al; 2023 | **[60]** | Case Control | Tunisia | PCR-SSP | AC | 81 | 100 | 2010 EULAR/ACR |
| **14** | Carthy et al; 1993 | **[61]** | Case Control | Greece | PCR-SSO | AF | 43 | 86 | 1956 ARA |
| **15** | Combe et al; 1995 | **[62]** | Case Control | France | PCR-SSO | AF | 94 | 489 | 1987 ACR |
| **16** | Cucchi-Mouillot et al; 1999 | **[63]** | Case Control | France | Reverse Hybridization | AF | 31 | 39 | 1987 ACR |
| **17** | de Juan et al; 1994 | **[64]** | Case Control | Spain | PCR-SSO | AF | 62 | 100 | 1987 ACR |
| **18** | Dhaouadi et al; 2010 | **[65]** | Case Control | Tunisia | PCR-SSP | AF | 90 | 100 | 1987 ACR |
| **19** | Djidjik et al; 2014 | **[66]** | Case Control | Algeria | PCR-SSP | AC | 134 | 132 | 1987 ACR |
| **20** | Farouk et al; 2009 | **[67]** | Case Control | Egypt | PCR-SSO | AC | 29 | 15 | 1987 ACR |
| **21** | Fathi N et al; 2008 | **[68]** | Case Control | Egypt | PCR-SSO | AC | 60 | 100 | 1987 ACR |
| **22** | Fathi S et al; 2010 | **[46]** | Case Control | Egypt | PCR-SSO | AF | 38 | 561 | 1987 ACR |
| **23** | Fejzic et al; 2017 | **[69]** | Case Control | Bosnia & Herzegovina | PCR-SSP | AC | 48 | 104 | 1987 ACR |
| **24** | Gonzalez-Escribano et al; 1999 | **[70]** | Prospective | Spain | PCR-SSO | AC | 137 | 305 | 1987 ACR |
| **25** | Hajeer et al; 2000 | **[71]** | Case Control | Spain | RDB Hybridization | AF | 179 | 145 | 1987 ACR |
| **26** | Ioannidis et al; 2002 | **[17]** | Case Control | Greece | PCR-SSO & PCR-SSP | AC | 174 | 103 | 1987 ACR |
| **27** | Kazkaz et al; 2007 (F) | **[47]** | Case Control | France | PCR-SSP | AC | 512 | 471 | 1987 ACR |
| **28** | Kazkaz et al; 2007 (S) | **[47]** | Case Control | Syria | PCR-SSP | AC | 156 | 120 | 1987 ACR |
| **29** | Kinikli et al; 2003 | **[72]** | Case Control | Turkey | PCR-SSP | AC | 104 | 100 | 1987 ACR |
| **30** | Klimenta et al; 2019 | **[73]** | Case Control | Bosnia & Herzegovina | PCR-SSP | AC | 83 | 82 | Clinical Assessment |
| **31** | Lagha et al; 2016 | **[44]** | Case Control | Tunisia | PCR-SSP | AC | 110 | 116 | 1987 ACR |
| **32** | Lopez-Arbesu et al; 2007 | **[74]** | Case Control | Spain | PCR-SSO & PCR-SSP | AC | 140 | 154 | 1987 ACR |
| **33** | Marinovic et al; 2022 (SR) | **[48]** | Case Control | Croatia | PCR-SSO & PCR-SSP | AC | 74 | 80 | 2010 EULAR/ACR |
| **34** | Marinovic et al; 2022 (SD) | **[48]** | Case Control | Croatia | PCR-SSO & PCR-SSP | AC | 74 | 80 | 2010 EULAR/ACR |
| **35** | Mourad et al; 2013 | **[75]** | Case Control | Syria | PCR-SSP | AC | 86 | 200 | 1987 ACR |
| **36** | Pascual et al; 2001 | **[76]** | Case Control | Spain | PCR-SSO | AC | 160 | 153 | 1987 ACR |
| **37** | Perdriger et al; 1996 | **[77]** | Case Control | France | RDB Hybridization | AC | 155 | 150 | 1987 ACR |
| **38** | Prifti-Kurti et al; 2014 | **[78]** | Case Control | Albania | PCR-SSP | AF | 100 | 191 | 1987 ACR |
| **39** | Reviron et al; 2001 | **[79]** | Case Control | France | PCR-SSO | AC | 203 | 230 | 1987 ACR |
| **40** | Said et al; 2021 | **[80]** | Case Control | Egypt | PCR-SSO | AC | 150 | 150 | Clinical Assessment |
| **41** | Salvarani et al; 1998 | **[81]** | Case Control | Italy | PCR-SSP | AF | 86 | 396 | 1987 ACR |
| **42** | Samy et al; 2000 | **[82]** | Case Control | Egypt | PCR-SSO | AC | 26 | 14 | 1987 ACR |
| 43 | Saruhan-Direskeneli et al; 1998 | [83] | Case Control | Turkey | PCR-SSO | AC | 101 | 101 | 1987 ACR |
| 44 | Sfar et al; 2010 | [84] | Case Control | Tunisia | PCR-SSP | AC | 132 | 110 | 1987 ACR |
| 45 | Soliman et al; 2016 | [85] | Case Control | Egypt | PCR-SSP | AC | 40 | 20 | 2010 EULAR/ACR |
| 46 | Stavropoulos et al; 1997 | [86] | Case Control | Greece | PCR-SSO | AC | 86 | 130 | 1987 ACR |
| 47 | Toussirot et al; 1998 | [87] | Case Control | France | PCR-SSO | AC | 104 | 74 | 1987 ACR |
| 48 | Ucar et al; 2012 | [88] | Case Control | Turkey | PCR-SSP | AC | 320 | 360 | 1987 ACR |
| 49 | Valenzuela et al; 1999 | [89] | Case Control | Spain | PCR-SSO | AC | 82 | 200 | 1987 ACR |
| 50 | Yelamos et al; 1993 | [90] | Case Control | Spain | PCR-SSO | AC | 70 | 189 | 1987 ACR |

HLA: Human Leukocyte Antigen; AC: Allele Count; AF: Allele Frequency; PCR-SSP: Polymerase Chain Reaction - Single Specific Primer, PCR-SSO: Polymerase Chain Reaction - sequence-specific oligonucleotide; RDB: Reverse dot-blot; ACR: American College of Rheumatology; EULAR: European League Against Rheumatism; ARA: American Rheumatism Association; SR: Sinj Region in Croatia; SD: Split-Dalmatia County in Croatia.

The allele frequencies for RA patients and controls, as well as individual study association results, are available upon request.

### Risk of bias

Risk of bias was assessed using the ROBINS-E tool across 50 populations from 48 studies, revealing variability in methodological quality. Thirteen studies were rated as low risk, while eight were classified as having some concerns, primarily due to issues related to confounding or missing data.

Twelve studies were classified as high risk, and sixteen were rated as very high risk, with uncontrolled confounding being the predominant concern. In addition, two studies were rated as high risk or as having some concerns specifically due to unreported diagnostic criteria for rheumatoid arthritis (Domain 6: outcome measurement bias).

Despite these concerns, exposure and outcome measurements were generally reliable, and selective reporting was not identified as a major source of bias. The ROBINS-E assessment is visually presented in the traffic light plot and summary plot (**Fig 3**).

**Fig 3:**
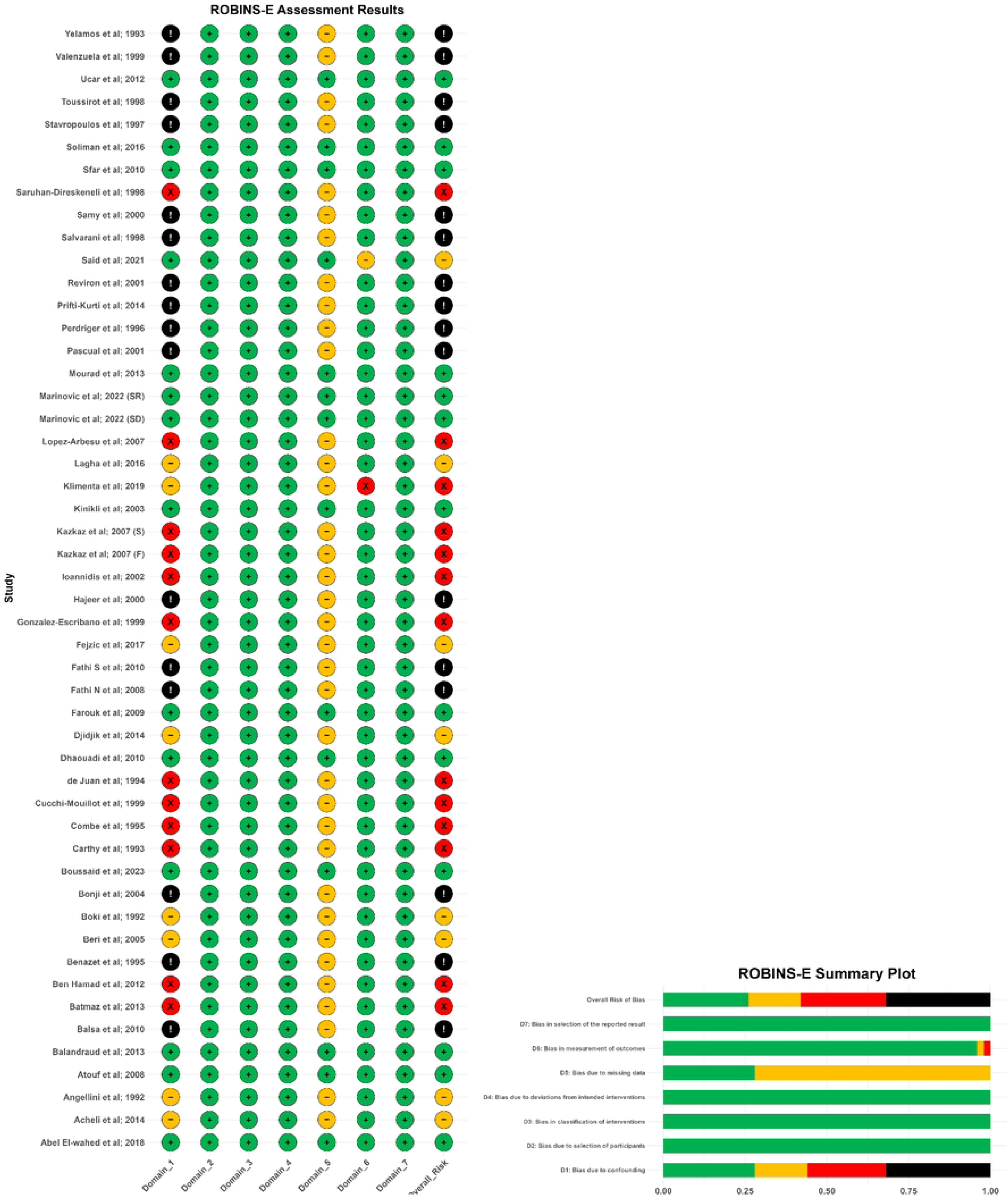
Risk of bias assessment in observational studies using ROBINS-E tool

### Outcome measures and statistical analysis

#### Association of *HLA-DRB1* alleles with rheumatoid arthritis

Our meta-analysis evaluated the association between *HLA-DRB1* alleles and RA risk in Mediterranean populations. Among the 13 analyzed alleles, four (*01, *04, *09, and *10) were significantly associated with RA susceptibility. *HLA-DRB1*\*04 was associated with the highest increased risk (OR 2.412, 95% CI [2.204–2.641]; *p* < 0.001), followed by *HLA-DRB1*\*10 (OR 1.649, 95% CI [1.419–1.917]; *p* < 0.001).

Conversely, seven alleles (*03, *07, *08, *11, *12, *13, and *14) were associated with a reduced RA risk. Among these, *HLA-DRB1*\*13 had the most pronounced inverse association (OR 0.580, 95% CI [0.527–0.638]; *p* < 0.001), closely followed by *HLA-DRB1*\*07 (OR 0.585, 95% CI [0.522–0.656]; *p* < 0.001).

Full statistical details are provided in **Table 2**, with forest plots available in (**S5 Appendix**).

**Table 2:** Key meta-analysis indicators of *HLA-DRB1* variations and rheumatoid arthritis susceptibility.

| *HLA Allele* | Studies | RA | Controls | Model Effect | OR | 95% CI | z | P-value | Test for heterogeneity |  |  |  |  |
| --- | --- | --- | --- | --- | --- | --- | --- | --- | --- | --- | --- | --- | --- |
|  |  |  |  |  |  |  |  |  | Q | df | p-value | I<sup>2</sup>(%) | PI |
| ***DRB1\*01*** | 41 | 1299/9882 | 1573/17988 | Random | 1.649 | 1.419 - 1.917 | 6.53 | **<0.001** | 102.94 | 40 | <0.001 | 61.1 | 0.792 - 3.434 |
| *DRB1\*01:01* | 14 | 381/3394 | 309/4574 | Random | 1.703 | 1.366 - 2.121 | 4.74 | **<0.001** | 21.73 | 13 | 0.06 | 40.2 | 0.919 - 3.151 |
| *DRB1\*01:02* | 11 | 73/2978 | 69/4128 | Random | 1.385 | 0.97 - 1.976 | 1.79 | 0.073 | 7.86 | 10 | 0.643 | 0 | 0.919 - 2.088 |
| *DRB1\*01:03* | 9 | 25/2476 | 69/3868 | Random | 0.737 | 0.407 - 1.337 | -1.00 | 0.316 | 10.05 | 8 | 0.261 | 20.4 | 0.223 - 2.443 |
| ***DRB1\*03*** | 38 | 848/9402 | 2014/16810 | Random | 0.823 | 0.718 - 0.943 | -2.80 | **0.005** | 70.33 | 37 | <0.001 | 47.4 | 0.459 - 1.476 |
| ***DRB1\*04*** | 43 | 2396/10040 | 2108/18284 | Random | 2.412 | 2.204 - 2.641 | 19.09 | **<0.001** | 57.16 | 42 | 0.059 | 26.5 | 1.777 - 3.274 |
| *DRB1\*04:01* | 24 | 864/8062 | 466/12350 | Random | 3.163 | 2.634 - 3.798 | 12.34 | **<0.001** | 33.31 | 23 | 0.0758 | 31 | 1.914 - 5.226 |
| *DRB1\*04:02* | 20 | 107/6328 | 154/10866 | Random | 0.915 | 0.626 - 1.336 | -0.46 | 0.645 | 30.03 | 19 | 0.051 | 36.7 | 0.306 - 2.733 |
| *DRB1\*04:03* | 18 | 80/5510 | 181/10088 | Random | 0.689 | 0.485 - 0.980 | -2.07 | **0.038** | 22.43 | 17 | 0.17 | 24.2 | 0.296 - 1.604 |
| *DRB1\*04:04* | 21 | 360/6620 | 248/10968 | Random | 2.647 | 2.228 - 3.146 | 11.06 | **<0.001** | 15.11 | 20 | 0.770 | 0 | 2.202 - 3.183 |
| *DRB1\*04:05* | 23 | 349/6806 | 171/11246 | Random | 2.967 | 2.197 - 4.006 | 7.10 | **<0.001** | 41.36 | 22 | 0.0075 | 46.8 | 1.079 - 8.158 |
| *DRB1\*04:06* | 9 | 10/3740 | 35/7240 | Random | 0.370 | 0.227 - 1.015 | -1.92 | 0.055 | 6.48 | 8 | 0.5934 | 0 | 0.195 - 1.184 |
| *DRB1\*04:07* | 14 | 42/4936 | 73/9288 | Random | 1.097 | 0.613 - 1.964 | 0.31 | 0.756 | 16.45 | 13 | 0.226 | 21.0 | 0.317 - 3.798 |
| *DRB1\*04:08* | 19 | 95/6128 | 38/10336 | Random | 2.847 | 1.605 - 5.051 | 3.58 | **<0.001** | 29.00 | 18 | 0.048 | 37.9 | 0.554 - 14.619 |
| ***DRB1\*07*** | 36 | 681/8748 | 1943/16104 | Random | 0.585 | 0.522 - 0.656 | -9.21 | **<0.001** | 41.17 | 35 | 0.219 | 15 | 0.440 - 0.777 |
| ***DRB1\*08*** | 36 | 182/8880 | 431/16244 | Random | 0.772 | 0.611 - 0.975 | -2.17 | **0.0298** | 45.46 | 35 | 0.111 | 23 | 0.382 - 1.559 |
| ***DRB1\*09*** | 30 | 119/7954 | 127/12918 | Random | 1.379 | 1.007 - 1.889 | 2 | **0.045** | 32.53 | 29 | 0.297 | 10.9 | 0.716 - 2.657 |
| ***DRB1\*10*** | 39 | 350/9540 | 325/17376 | Random | 2.034 | 1.659 - 2.493 | 6.83 | **<0.001** | 46.98 | 38 | 0.151 | 19.1 | 1.137 - 3.638 |
| ***DRB1\*10:01*** | 11 | 139/3540 | 59/3396 | Random | 2.275 | 1.532 - 3.377 | 4.08 | **<0.001** | 13.46 | 10 | 0.199 | 25.7 | 0.950 – 5.448 |
| ***DRB1\*11*** | 36 | 896/8574 | 2306/15982 | Random | 0.634 | 0.599 - 0.713 | -9.63 | **<0.001** | 32.96 | 35 | 0.557 | 0 | 0.598 - 0.715 |
| ***DRB1\*12*** | 33 | 99/8206 | 281/15586 | Random | 0.696 | 0.544 - 0.890 | -2.89 | **0.0039** | 25.32 | 32 | 0.793 | 0 | 0.539 – 0.899 |
| ***DRB1\*13*** | 35 | 667/8390 | 2104/15814 | Random | 0.580 | 0.527 - 0.638 | -11.13 | **<0.001** | 25.18 | 34 | 0.864 | 0 | 0.525 - 0.640 |
| ***DRB1\*14*** | 35 | 217/8390 | 621/15814 | Random | 0.716 | 0.605 - 0.849 | -3.86 | **<0.001** | 32.76 | 34 | 0.5284 | 0 | 0.601 - 0.854 |
| ***DRB1\*15*** | 32 | 745/7870 | 1422/14394 | Random | 0.940 | 0.845 - 1.046 | -1.14 | 0.256 | 32.47 | 31 | 0.394 | 0 | 0.790 - 1.118 |
| ***DRB1\*16*** | 31 | 337/7750 | 604/14194 | Random | 0.936 | 0.792 - 1.107 | -0.77 | 0.441 | 33.57 | 30 | 0.299 | 10.6 | 0.660 - 1.329 |

HLA: Human Leucocyte Antigen; OR: odds ratio; 95% CI: confidence interval at 95%, z: The z-value tests, P-value: Significance level of the association. $P < 0.05$ indicates statistical significance, Q: Cochran's Q statistic, df: Degrees of freedom, $I^2$ : Inconsistency test, PI: Prediction Intervals.

#### Association of *HLA-DRB1* subgroups with rheumatoid arthritis

Further analysis of *HLA-DRB1*\*01, *04, and *10 subgroups revealed significant associations with RA susceptibility. Six subgroup alleles (*01:01, *04:01, *04:04, *04:05, *04:08, and *10:01) correlated strongly with RA risk. *HLA-DRB1*\*04:01 was prominently associated with increased RA risk (OR 3.163, 95% CI [2.634–3.798]; *p* < 0.001), followed by *HLA-DRB1*\*04:05 (OR 2.967, 95% CI [2.197–4.006]; *p* < 0.001). Among the subtypes analyzed, *HLA-DRB1*\*04:03 emerged as more common in controls (OR 0.689, 95% CI [0.485–0.980]; p = 0.038). Detailed results for all subgroups are available in **Table 2**, with corresponding forest plots in (**S5 Appendix**).

#### Regional differences in *HLA-DRB1* associations with rheumatoid arthritis

To evaluate the influence of geographic region on *HLA-DRB1* associations with RA, a subgroup meta-analysis was performed, stratifying studies into Southern European and MENA populations. The overall patterns of association were comparable across both regions, with no statistically significant subgroup differences observed for any allele. Tests for subgroup differences yielded χ² values (df = 1) ranging from < 0.001 to 3.24, with corresponding *p*-values between 0.07 and 0.97 (all > 0.05). Full forest plots are available in (**S6 Appendix**).

#### Overall risk of bias for each meta-analysis

Out of the 13 *HLA-DRB1* allele-level meta-analyses, only *HLA-DRB1*\*01 was assessed as having a high overall risk of bias; the remaining 12 were rated as presenting some concerns. In the subgroup-level analyses, the distribution of overall risk of bias was as follows: three meta-analyses were rated as having some concerns, seven as high risk, and two as very high risk. A detailed visual summary of these findings is provided in the corresponding forest plots. (S1 ***Checklist-PRISMA 2020 Checklist***

**S2 Checklist-PRISMA Abstract Checklist**

**S3 Checklist-Genetic Association Meta-analysis Checklist S4 Appendix**).

### Heterogeneity analysis

Heterogeneity was assessed using the Q-statistic, I², and Prediction Intervals (PIs), with the primary results presented in **Table 2**. Significant heterogeneity (Q-statistic *p* < 0.05, I² = 37.9% – 61.1%) was observed for *HLA-DRB1*\*01, *03, and *04:08. Moreover, these alleles included the null value (OR=1) within their 95% PIs.

Additionally, *HLA-DRB1*\*04:03, *07, *08, *09, and *1001 yielded lower heterogeneity levels (I² = 10.9% – 40.2%, Q-statistic *p* > 0.05) but also included the null value within their 95% PIs.

Meta-regression between Southern Europe and MENA populations did not reveal significant differences (**S7 Appendix**).

Outlier removal effectively reduced heterogeneity for *HLA-DRB1*\*01, lowering I² to 13% (Q-statistic *p* = 0.25) and narrowing its 95% PI to [1.2, 1.97]. However, heterogeneity persisted for other alleles despite similar corrections (**S8 Appendix**). For alleles where heterogeneity remained after outlier removal, the percentages of true effect sizes expected to fall below or above an odds ratio of 1 were estimated using 95% PIs and the cumulative distribution function (CDF). These results, provided in (**Fig 4**) and **Table 3**, offer a detailed account of the variability across studies.

**Fig 4:**
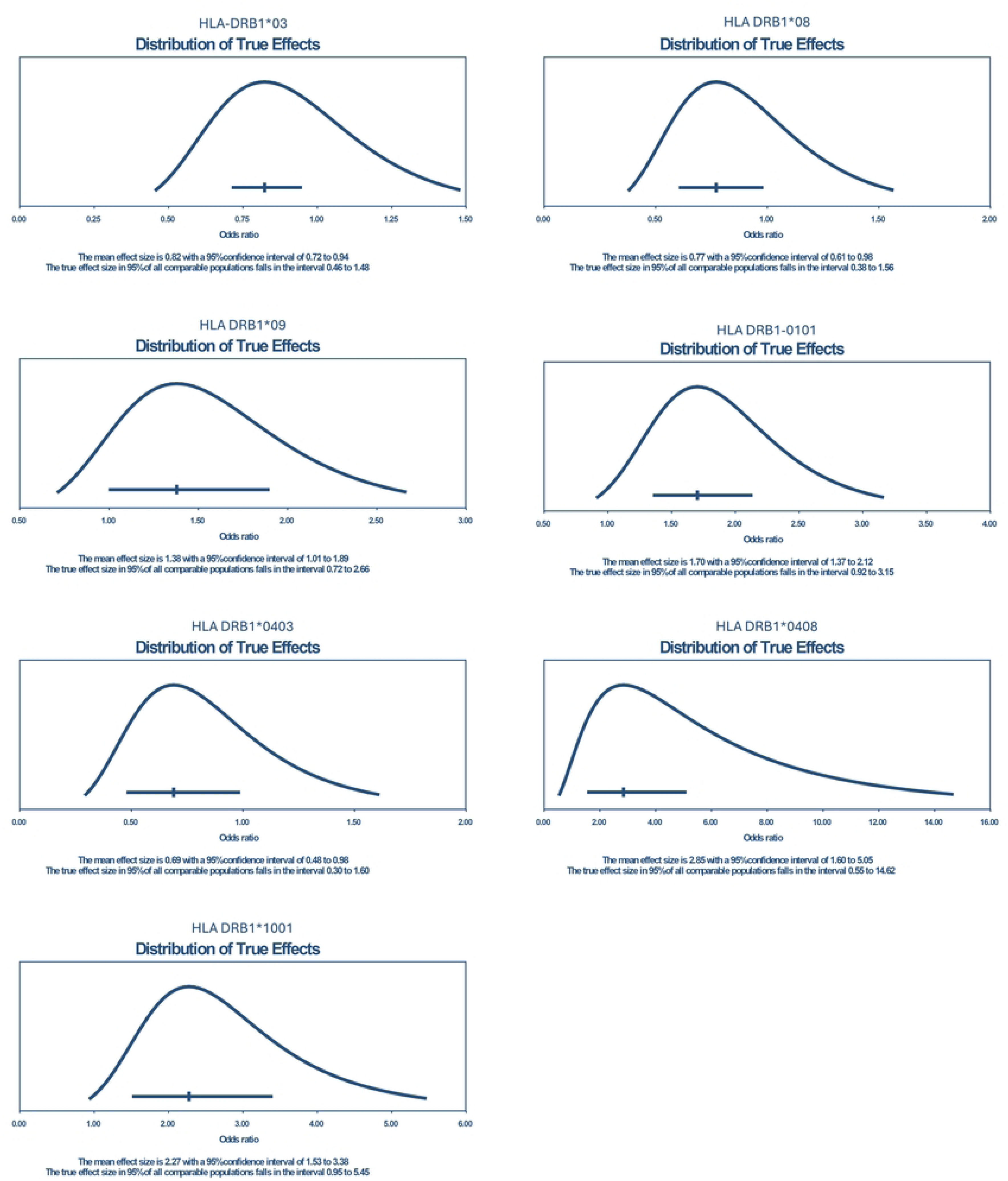
Presents the distribution of true effects for selected *HLA-DRB1* alleles, illustrating the range of plausible effect sizes across comparable populations. Each curve represents the estimated distribution of the true odds ratio (OR) values, with the mean effect size and its 95% confidence interval (CI) indicated below each plot. The prediction interval (PI), displayed as a horizontal line, represents the range within which the true effect size is expected to fall in 95% of comparable populations.

**Table 3:** Heterogeneity Metrics for *HLA-DRB1* Alleles.

| Allele | OR | $I^2$ (%) | 95% PI | Percentage of Comparable populations within 95%PI Range |
| --- | --- | --- | --- | --- |
| *HLA-DRB1* *03 | 0.823 | 47.4 | 0.459 - 1.476 | 74.37% < 1, 25.63% > 1 |
| *HLA-DRB1* *08 | 0.772 | 23 | 0.382 - 1.559 | 76.50% < 1, 23.50% > 1 |
| *HLA-DRB1* *09 | 1.379 | 10.9 | 0.716 - 2.657 | 16.84% < 1, 83.16% > 1 |
| *HLA-DRB1* *01:01 | 1.703 | 40.2 | 0.919 - 3.151 | 04.51% < 1, 95.49% > 1 |
| *HLA-DRB1* *04:03 | 0.689 | 24.1 | 0.300 - 1.600 | 80.60% < 1, 19.40% > 1 |
| *HLA-DRB1* *04:08 | 2.847 | 37.9 | 0.554 - 14.62 | 10.51% < 1, 89.49% > 1 |
| *HLA-DRB1* *10:01 | 2.275 | 25.7 | 0.950 – 5.448 | 03.26% < 1, 96.74% > 1 |

*HLA*: Human Leucocyte Antigen; *OR*: odds ratio; 95% *CI*: confidence interval at 95%, $I^2$ : Inconsistency test, *PI*: Prediction Intervals

*HLA-DRB1*\*04:05 was characterized by considerable heterogeneity (Q-statistic p < 0.05, I² = 46.8%), with a 95% PI of [1.079 – 8.158].

### Publication bias

The potential impact of publication bias was assessed using the trim-and-fill method. Based on predefined classification criteria, minimal publication bias was observed for *HLA-DRB1*\*01, *04, *10, *12, *14, *15, *01:02, *01:03, *04:03, *04:04, *04:07, and *10:01. Moderate bias was found for *HLA-DRB1*\*03, *07, *08, *13, and *04:05, while substantial bias was evident for *HLA-DRB1*\*09, *16, *04:01, and *04:08. No publication bias was detected for *HLA-DRB1*\*01:01 and *04:02. Full imputation results and funnel plots are provided in (**S9 Appendix**).

Trim and fill method was not applicable for *HLA-DRB1*\*11 and *04:06, as their effect size distribution was symmetrical in forest plots (**S9 Appendix**) and their I^2^= 0. This symmetry was further confirmed by Rücker’s test, with p-values of 0.256 and 0.146, respectively (both > 0.1) indicating no publication bias.

### Sensitivity analysis

#### Removal of very high risk of bias studies

Removal of studies rated as having a very high risk of bias yielded consistent results for most HLA-DRB1 alleles, with confidence intervals overlapping those of the primary analysis (**S10 Appendix**). *HLA-DRB1*\*04 and *10 retained their strong risk associations (OR = 2.48, 95% CI [2.22–2.77] and OR = 2.07, 95% CI [1.62–2.65], respectively), while *HLA-DRB1*\*13 continued to show the lowest odds ratio among protective alleles (OR = 0.57, 95% CI [0.51–0.63]). However, *HLA-DRB1*\*03 and *09 were no longer statistically significant following exclusion of high-risk studies. *HLA-DRB1*\*03 had an OR = 0.88 (95% CI [0.74–1.04], *p* = 0.12) and *HLA-DRB1*\*09 had an OR = 1.38 (95% CI [0.94–2.03], *p* = 0.10). The protective effects of *HLA-DRB1*\*07, *08, *11, *12, and 14 were maintained, while *HLA-DRB1*\*15 and *16 remained non-significant.

#### Leave one out analysis

For most *HLA-DRB1* alleles and subtypes, removal of individual studies had no meaningful impact on the pooled effect sizes, and statistical significance was retained. However, *HLA-DRB1*\*08, 09, and 04:03 lost statistical significance when specific studies were excluded. Detailed results are shown in the leave-one-out forest plots (**S11 Appendix**).

#### Trim-and-fill analysis

Following trim-and-fill adjustment, most *HLA-DRB1* alleles showed minimal change in effect estimates, and statistical significance remained consistent with the primary results. *HLA-DRB1*\*08 lost statistical significance after imputation, while *HLA-DRB1*\*01:02 reached statistical significance post-adjustment. For the remaining alleles, no changes in statistical significance were observed. Corresponding forest plots are presented in (**S12 Appendix**).

### Certainty of evidence assessment

Certainty of evidence for each *HLA-DRB1* allele was evaluated using the GRADE framework. All included studies lacked sufficient adjustment for prespecified confounding factors; therefore, no upgrading was applied. Across alleles, certainty ratings ranged from low to very low, based on domain-level assessments of risk of bias, inconsistency, and publication bias. Final GRADE ratings are summarized in **Table 4**.

**Table 4:** GRADE assessment for certainty of evidence:

| Outcome | Risk of Bias | Inconsistency | Imprecision | Indirectness | Publication Bias | Upgrading Factors | Overall Certainty |
| --- | --- | --- | --- | --- | --- | --- | --- |
| *HLA-DRB1\*01* and RA risk | Serious | Serious | None | None | Low | None | **Very Low** |
| *HLA-DRB1\*03* and RA protection | Moderate | Serious | 95% CI Close to Null* | None | Moderate | None | **Very Low** |
| *HLA-DRB1\*04* and RA risk | Moderate | Low | None | None | Moderate | Large Effect Size | **Low** |
| *HLA-DRB1\*07* and RA protection | Moderate | Low | None | None | Serious | None | **Very Low** |
| *HLA-DRB1\*08* and RA protection | Moderate | Low | 95% CI Close to Null* | None | Moderate | None | **Very Low** |
| *HLA-DRB1\*09* and RA risk | Moderate | Low | 95% CI Close to Null* | None | Very serious | None | **Very Low** |
| *HLA-DRB1\*10* and RA risk | Moderate | Low | None | None | Low | Large Effect Size | **Low** |
| *HLA-DRB1\*11* and RA protection | Moderate | Low | None | None | None | None | **Very Low** |
| *HLA-DRB1\*12* and RA protection | Moderate | Low | 95% CI Close to Null* | None | Moderate | None | **Very Low** |
| *HLA-DRB1\*13* and RA protection | Moderate | Low | None | None | Moderate | None | **Very Low** |
| *HLA-DRB1\*14* and RA protection | Moderate | Low | None | None | Low | None | **Very Low** |
| *HLA-DRB1\*01:01* and RA risk | Serious | Moderate | None | None | Low | None | **Very Low** |
| *HLA-DRB1\*04:01* and RA risk | Serious | Moderate | None | None | Very serious | Large Effect Size | **Very Low** |
| *HLA-DRB1\*04:03* and RA protection | Serious | Low | None | None | Low | None | **Very Low** |
| *HLA-DRB1\*04:04* and RA risk | Moderate | Low | None | None | Moderate | Large Effect Size | **Low** |
| *HLA-DRB1\*04:05* and RA risk | Serious | Moderate | None | None | Moderate | Large Effect Size | **Very Low** |
| *HLA-DRB1\*04:08* and RA risk | Serious | Moderate | None | None | Very serious | Large Effect Size | **Very Low** |
| *HLA-DRB1\*10:01* and RA risk | Serious | Moderate | None | None | Low | Large Effect Size | **Low** |

\*The "null" value refers to an Odds Ratio (OR) of 1, HLA: Human leucocyte antigen, RA: Rheumatoid arthritis, $I^2$ : Inconsistency test, CI: confidence interval

### Post hoc power analysis

Statistical power estimates were computed for each *HLA-DRB1* allele meta-analysis comparison. Most alleles demonstrated high power (≥ 0.80). Power estimates, along with allele-specific proportions and effect sizes, are presented in (**S13 Table**).

## Discussion

This meta-analysis provides a comprehensive evaluation of the association between *HLA-DRB1* alleles and RA susceptibility and protection in Mediterranean populations. By integrating advanced statistical approaches, this study refines risk estimates while accounting for variability across studies. The use of meta-regression, prediction intervals, and cumulative distribution function analysis allows for a more precise assessment of allele-specific associations and potential sources of heterogeneity. Additionally, the systematic evaluation of bias using the ROBINS-E framework enhances the reliability of the findings, while the GRADE approach provides a transparent assessment of the overall quality of evidence. These methodological considerations strengthen the validity of our conclusions and will be further explored in the discussion.

### *HLA-DRB1* association with RA and comparison with prior research

This meta-analysis identified multiple *HLA-DRB1* alleles that contribute to RA susceptibility within Mediterranean populations*. HLA-DRB1*\*01, *04, *09, and *10 emerged as key risk alleles, with *04 and 10 displaying the most pronounced effect sizes supporting their well-established involvement in RA pathogenesis through shared epitope driven mechanisms. Several sub-alleles (*HLA-DRB1*\*01:01, *04:01, *04:04, *04:05, 04:08, and 10:01) were likewise implicated in increased disease risk.

These findings align with previous meta-analyses conducted in diverse populations. For instance, studies of Arab populations identified *HLA-DRB1*\*04 and *HLA-DRB1*\*10 as crucial risk factors for RA [45]. Similar associations were observed in Asian populations, where *HLA-DRB1*\*04, *HLA-DRB1*\*04:01, *HLA-DRB1*\*04:05, and *HLA-DRB1*\*04:10 significantly elevated RA risk [91, 92]. Research on Chinese populations confirmed that *HLA-DRB1*\*04, *HLA-DRB1*\*04:01, *HLA-DRB1*\*04:04, *HLA-DRB1*\*04:05, and *HLA-DRB1*\*04:10 were associated with increased RA susceptibility [93]. In European populations, studies highlighted *HLA-DRB1*\*04, *HLA-DRB1*\*01:01, *HLA-DRB1*\*01:02, and *HLA-DRB1*\*10 as significant risk alleles [17, 94]. Similarly, in Latin American populations, Delgado-Vega et al. (2007) reported that *HLA-DRB1*\*04 and SE alleles were significant risk factors for RA [95]. These recurrent results across diverse ethnic groups underscore the pivotal role of SE alleles in RA pathogenesis.

Importantly, our meta-analysis is the first to show a significant association between *HLA-DRB1*\*09 and RA risk in Mediterranean populations, an association not previously observed in similar meta-analyses of European [94], Mediterranean European [17], Arabian [45], Asian [92], Asian-Mongoloid [91], Chinese [93], Latin American [95], Native American [96], or multi-ethnic [97, 98] populations. *HLA-DRB1*\*09 has been shown to correlate with RA risk in Malaysia [99], Korea [100, 101], Turkey [88], and Japan [102]. Further research is needed to explore how regional factors, such as diet, infection exposure, and cultural practices, interact with *HLA-DRB1*\*09 to influence RA risk.

### Mechanisms of susceptibility and pathogenesis

The association of *HLA-DRB1*\*01, *04, *09, and *10 with RA in this study reflects distinct but convergent mechanisms related to antigen presentation, T-cell activation, and inflammatory cytokine signaling. The *HLA-DRB1* SE alleles (*01, *04, and *10) are well-documented in RA pathogenesis due to their ability to bind and present citrullinated peptides, leading to the activation of autoreactive CD4+ T cells and the production of anti-citrullinated protein antibodies (ACPA), which are key drivers of disease progression [8, 103]. These alleles share a conserved amino acid sequence at positions 70–74 of the *HLA-DRβ* chain, which enhances their affinity for citrullinated antigens, thereby shaping a proinflammatory immune response characterized by the expansion of Th1 and Th17 cells and elevated production of IL-6 and TNF-α [104].

On the other hand, *HLA-DRB1*\*09:01, despite lacking the SE motif, remains strongly associated with RA, likely through an alternative antigen presentation pathway that does not depend on citrullination [105]. The presence of glutamic acid at position 74 in *09:01 alters its peptide-binding properties, potentially favoring non-citrullinated or uniquely modified antigens, which may explain its stronger association with seronegative RA [11, 106].

Moreover, *HLA-DRB1*\*04:05 and *10:01 exhibit a strong interaction with smoking, a key environmental risk factor, further amplifying the inflammatory response and accelerating joint damage [107, 108]. Interestingly, *09:01 does not independently interact with smoking but, when co-inherited with SE alleles, contributes to a synergistic effect, particularly in ACPA-negative RA cases, reinforcing the notion that multiple immune pathways contribute to disease susceptibility [105].

In contrast, Lagutkin et al 2025, provided strong evidence that *HLA-DRB1*\*09 and *15 contribute to ACPA production independently of the shared epitope [109]. These findings suggest that *HLA-DRB1*\*01, *04, *09, and *10 influence RA through both citrulline-dependent and citrulline-independent mechanisms, ultimately converging on pathways that drive chronic synovial inflammation, autoimmunity, and joint destruction.

### *HLA-DRB1* protection against RA and comparison with prior research

Seven *HLA-DRB1* alleles (*03, *07, *08, *11, *12, *13, and *14) were observed at significantly lower frequencies among RA cases than controls, indicating a protective association. Among these, *HLA-DRB1*\*13 and *07 exhibited the strongest and most enduring protective effects across studies. In terms of subgroups, *HLA-DRB1*\*04:03 was the only variant associated with a reduced risk of RA.

These findings align with previous research. In Arab populations, Bizzari et al. (2017) reported that *HLA-DRB1*\*03, *HLA-DRB1*\*07, *HLA-DRB1*\*11, and *HLA-DRB1*\*13 conferred protection against RA [45]. Similarly, European studies identified *HLA-DRB1*\*13 as a protective allele [94]. Chinese studies also demonstrated protective associations for *HLA-DRB1*\*11, *HLA-DRB1*\*13, and *HLA-DRB1*\*14 [93]. A recent multi-ethnic study also identified *HLA-DRB1*\*03, *HLA-DRB1*\*07, and *HLA-DRB1*\*13 as protective alleles [98].

Notably, the majority of these alleles lack the shared epitope motif, supporting the hypothesis that the absence of this sequence or the presence of alternative motifs such as the DERAA sequence found in certain HLA-DRB1 alleles may confer reduced susceptibility to RA [12]. The broadly replicated protective associations observed across diverse populations highlight the broad relevance of these HLA-DRB1 alleles in reducing RA susceptibility, underscoring the critical role of genetic factors in the pathogenesis and potential stratification of rheumatoid arthritis [110].

### Regional comparison: geographic variability in *HLA-DRB1* associations

The regional subgroup meta-analysis, comparing Southern European and MENA populations, revealed no significant differences in the associations of *HLA-DRB1* alleles with RA risk. This pattern suggests that despite potential variations in environmental exposures and genetic backgrounds, the genetic impact of *HLA-DRB1* alleles on RA susceptibility remains conserved across Mediterranean populations. These findings corroborate with the historical genetic continuity of Mediterranean populations, which share common ancestry due to migration, trade, and admixture over centuries [111].

### Risk of bias in included studies

The ROBINS-E evaluation highlighted substantial methodological limitations across the included studies, particularly due to uncontrolled confounding, missing data, and inconsistent RA diagnostic criteria. Confounding emerged as the most influential source of bias, with many studies failing to adjust for key variables such as age, sex, geographic origin, and environmental exposures; factors that may distort allele–disease associations [22]. Although exposure and outcome assessments were generally sound, the cumulative impact of bias across studies varied, especially among subgroup analyses. These findings underscore the need for stricter methodological standards, improved confounder adjustment, and diagnostic harmonization to strengthen the validity of genetic association studies in RA.

### Addressing heterogeneity

The analysis revealed substantial heterogeneity across Mediterranean populations in the association between *HLA-DRB1* alleles and RA, with variability in effect sizes quantified using I² and Q-statistics. While some alleles (*HLA-DRB1*\*01, *03, and *04:08) were marked moderate-to-high heterogeneity (I² ∼38–61%), reflecting variability in their association with RA, others (*HLA-DRB1*\*04:03, *07, *08, *09, and *10:01) displayed lower heterogeneity yet retained wide 95% prediction intervals, implying possible null effects in some populations. Conversely, *HLA-DRB1*\*11, *12, *13, and *14 alleles demonstrated uniform associations across studies (I² = 0%), highlighting the homogeneity of their protective effects against RA.

The meta-regression assessing regional differences (Southern Europe vs. MENA) found no statistically significant variation, implying that subregional factors did not substantially contribute to heterogeneity [29]. However, other variables, including study design, allele frequency shifts, and unmeasured environmental interactions, may have influenced the observed variability [1, 112].

Outlier screening helped isolate the source of dispersion in specific cases. For *01, the removal of a few studies altered the overall estimate and narrowed the prediction interval.

This effect was not replicated in other alleles, where adjustments had a minimal influence on the overall distribution of findings.

To further contextualize heterogeneity, cumulative frequency distributions were constructed from the 95% prediction intervals to characterize the expected distribution of effect sizes across comparable future populations. Unlike conventional meta-analyses that report a single pooled estimate, CFDs estimate the proportion of populations in which a given allele is likely to exert a risk or protective effect. For instance, odds ratios for HLA-DRB103, 07, and 04:03 were below 1 in 73–81% of comparable populations, whereas those for *09, *01:01, 04:08, and 10:01 exceeded 1 in 87–97% of cases. These proportions reflect how frequently each allele’s effect size would align with a risk-or protective-oriented direction if the analysis were repeated in similar populations. This approach does not estimate probability within the current data but describes the degree of directional consistency across comparable study settings [33].

*HLA-DRB1*\*04:05 showed considerable variation across studies, with I² around 47% and a prediction interval spanning 1.08 to 8.16. This range reflects differing effect estimates across populations, with some reporting relatively low risk estimates and others reporting much higher values. Such variation may be influenced by differences in linkage disequilibrium with nearby alleles, shifts in allele frequencies, or population-specific environmental exposures [11].

### Publication bias analysis

Assessment of publication bias indicated variable degrees of funnel plot asymmetry across *HLA-DRB1* alleles. *HLA-DRB1*\*01, *04, *10, *12, *14, and *15 showed minimal evidence of bias, with few imputed studies identified by the trim-and-fill method, lending greater confidence to their observed associations. In contrast, moderate asymmetry for *HLA-DRB1*\*03, *07, *08, *13, and *04:05 suggest a possible overestimation of their risk or protective effects, likely due to selective reporting of significant findings. Greater asymmetry was detected for *HLA-DRB1*\*09, *16, *04:01, and 04:08, with 6–8 potentially missing studies. This raises concern that the observed odds ratios for these alleles; particularly *HLA-DRB1*\*09; may be inflated, and that their true effect sizes are likely more modest than initially reported.

HLA-DRB1*01:01, *04:02, *11, and *04:06 showed no evidence of publication bias, strengthening confidence in the reliability of their observed associations. These results underscore the importance of cautious interpretation, as selective reporting can contribute to inflated estimates of genetic risk. To enhance the validity of future meta-analyses, approaches such as selection models or the inclusion of unpublished data should be considered to minimize bias and improve the credibility of findings [113, 114].

### Sensitivity analyses: Interpretation and implications

Excluding studies with a very high risk of bias did not alter the associations observed for *HLA-DRB1*\*04 and *10, both of which retained elevated odds ratios and statistical significance. Likewise, *HLA-DRB1*\*13, *07, *08, *11, *12, and *14 remained significantly associated with a reduced risk of RA, indicating that these findings were not dependent on the inclusion of low-quality data. In contrast, statistical significance was not retained for *HLA-DRB1*\*03 and *09 after excluding high-risk studies, raising the possibility that their observed associations were shaped by methodological limitations. This outcome highlights the extent to which pooled genetic estimates can be influenced by the quality of included evidence [115].

The leave-one-out analysis showed that the primary associations remained largely unaffected by the exclusion of individual studies. This finding strengthens the evidence for the associations involving *HLA-DRB1*\*04, *HLA-DRB1*\*10, and *HLA-DRB1*\*13 alleles. However, the change in significance for *HLA-DRB1*\*08, *09, and *04:03 following the removal of individual studies points to a dependence on specific datasets. This pattern calls for careful interpretation and highlights the importance of additional studies to clarify their associations with RA.

This finding exemplifies a broader methodological challenge in genetic association studies, wherein marginal or context-dependent signals are particularly susceptible to distortion from study-level heterogeneity and sampling variability, thereby complicating the interpretation of results and potentially leading to overestimated or spurious associations[116].

Adjustments for publication bias using the trim-and-fill method showed that most associations were unaffected; however, *HLA-DRB1*\*08 lost its protective significance, while *HLA-DRB1*\*01:02, a rare allele subtype, became statistically significant post-adjustment. These shifts point to the influence of selective reporting in shaping the apparent strength of associations. Routine use of funnel plots and correction techniques remains essential for evaluating the credibility of meta-analytic findings in genetic research [34].

### Quality of evidence

Although most allele-level comparisons exhibited sufficient post hoc statistical power, the certainty of evidence remained consistently low or very low across outcomes. Strong associations observed for *HLA-DRB1*\*04, *04:04, *10, and *10:01 were not upgraded in GRADE ratings due to unresolved confounding, selective reporting bias, and study design limitations. Protective alleles, including *HLA-DRB1*\*03, *07, *08, *11, *12, *13, and *14, were similarly rated as very low certainty, primarily reflecting imprecision and persistent risk of bias. Even *HLA-DRB1*\*01 and *09, despite achieving statistical significance with adequate power, were downgraded: *01 due to considerable heterogeneity and *09 due to evidence of publication bias and effect estimates clustering near the null [26].

These findings illustrate a fundamental challenge in genetic epidemiology: statistical significance and high power alone cannot compensate for underlying methodological weaknesses. Associations that appear statistically convincing may nonetheless rest on fragile evidence when risk of bias, inconsistency, and indirectness are systematically considered. Notably, even alleles exhibiting minimal heterogeneity, such as the protective association of *11, did not escape downgrading due to cumulative limitations across multiple GRADE domains [26, 113].

The total sample size (over 6,000 RA cases and 10,000 controls across 48 studies) was adequate to support the research objectives. Post hoc power analyses confirmed that most allele-level comparisons, including those associated with uncertainty in sensitivity analyses, such as *08 and *09, were adequately powered to detect meaningful effects. Yet the enduring instability seen in sensitivity points to deeper issues of residual bias and unmeasured confounding rather than sample size inadequacy.

Together, these findings highlight the need for future genetic association studies to place equal importance on study design quality and sample size. Prospective registration, bias mitigation strategies at the design stage, full transparency in reporting, and careful adjustment for environmental confounders will be essential to produce evidence that is both scientifically robust and clinically actionable in personalized RA management [117, 118].

### Limitations

This review employed established methodological tools and sensitivity strategies; however, several limitations remain. Study quality varied, with several studies showing high to very high risk of bias due to confounding factors. While sensitivity analyses were performed, residual biases may still influence some pooled estimates. Additionally, publication bias remains a concern, as some negative findings may not have been reported, potentially overemphasizing certain allele associations [119].

Several alleles displayed considerable heterogeneity, with effect estimates differing substantially between populations. While meta-regression and outlier analysis helped clarify part of this variation unaccounted factors such as gene-gene interactions and environmental exposures could still influence these findings [120]. Uneven population coverage further limits generalizability, as data from the Eastern Mediterranean and North Africa were scarce, making findings more reflective of well-studied regions like Spain, Farnce, and Egypt.

Hardy-Weinberg equilibrium (HWE) testing was not uniformly applied in this meta-analysis, as many of the included studies did not report genotype-level data for *HLA-DRB1* alleles or did not assess HWE in control groups. Since this review focused on allele-level rather than genotype-level associations, formal HWE testing could not be consistently conducted across datasets. Nevertheless, the absence of HWE verification may limit the ability to detect potential genotyping errors, selection bias, or population substructure in the source studies [121].

Finally, causality cannot be established, as *HLA-DRB1* alleles are in linkage disequilibrium with other HLA loci, therefore, the reported associations might reflect effects from adjacent causal variants rather than the studied alleles themselves [10]. Addressing these limitations will require future studies to incorporate broader population representation, utilize patient-level data, and apply high-resolution genotyping to clearly differentiate causal variants from proxy markers.

### Clinical implications

The findings of this study offer actionable insights for improving RA diagnosis and treatment strategies. Identifying *HLA-DRB1* alleles associated with susceptibility and protection may support the development of genetic screening programs aimed at identifying individuals at elevated risk. Such initiatives could enable earlier diagnosis and intervention, with the potential to slow disease progression and enhance clinical outcomes. In parallel, recent research indicates that carriers of shared epitope alleles, particularly those with valine at position 11, respond more favorably to abatacept than to TNF inhibitors [122, 123]. Incorporating genetic markers into treatment decision-making may help facilitate more personalized and effective therapeutic approaches.

### Future directions

Future research should adopt standardized study designs and ensure adequate control for confounding variables. Expanding datasets to include underrepresented Mediterranean regions would improve the generalizability of findings across diverse populations. Investigating gene–environment interactions, particularly between *HLA-DRB1* alleles and region-specific exposures, may clarify mechanisms of RA susceptibility and inform regionally relevant risk assessments.

Methodological improvements, such as larger sample sizes, consistent adjustment for confounders, and full reporting of both significant and non-significant findings, are essential to reduce bias in genetic association studies. Ongoing synthesis of emerging data through updated meta-analyses can improve the precision of effect estimates. Additional research is also warranted to determine whether RA-associated genetic markers have practical value in patient stratification or in guiding treatment decisions.

## Conclusions

This meta-analysis identified *HLA-DRB1*\*04 and 10 as the alleles most strongly associated with an increased risk of rheumatoid arthritis in Mediterranean populations, whereas HLA-*DRB1*\*13 and *07 were the most reliably associated with reduced risk. An additional association was observed for *HLA-DRB1*\*09, though this finding was affected by instability across sensitivity analyses and supported by very low certainty of evidence.

While several alleles showed considerable heterogeneity in effect size, comparisons between Southern European and MENA populations did not yield significant regional differences. Exclusion of studies with a very high risk of bias had limited influence on most pooled estimates.

Overall, the associations between *HLA-DRB1* alleles and rheumatoid arthritis were based on evidence graded as low or very low certainty. These findings call attention to the need for additional research using larger sample sizes, standardized study designs, proper adjustment for confounding variables, and measures to address publication bias. The repeated observation of specific allele patterns across studies points to the relevance of *HLA-DRB1* typing in risk stratification and provides a rationale for exploring geographically adapted genetic screening strategies.

## Data Availability

All data supporting the findings of this study are available within the manuscript and its Supporting Information files. Additional data and analysis code can be made available upon reasonable request to the corresponding author.

**S1 Checklist-PRISMA 2020 Checklist**

**S2 Checklist-PRISMA Abstract Checklist**

**S3 Checklist-Genetic Association Meta-analysis Checklist**

**S4 Table-List of excluded full-text studies with reasons**

**S5 Appendix-Forest Plots**

**S6 Appendix-Regional subgroup meta-analyses forest plots**

**S7 Appendix-Meta Regression Diagnostic Plots**

**S8 Appendix-Outliers Removed Forest Plots**

**S9 Appendix-Trim and Fill Funnel Plots**

**S10 Appendix-Sensitivity analyses-removal of very high ROB studies**

**S11 Appendix-Sensitivity analyses-Leave One Out**

**S12 Appendix-Sensitivity analyses-Trim and Fill**

**S13 Table-Post hoc power analysis for each *HLA-DRB1* allele meta-analysis**

